# ASSOCIATION OF ANEMIA WITH HEALTH-RELATED QUALITY OF LIFE AMONG HIV-INFECTED INDIVIDUALS IN MERU, KENYA

**DOI:** 10.1101/2024.07.23.24310855

**Authors:** Nasteha Adan Issack, Elizabeth Kamolo, Joan J Simam, Frank G. Onyambu

**Affiliations:** Meru University of Science and Technology, Meru, Kenya; Centre for Molecular Bioinformatics and Genomics, Nairobi, Kenya

**Keywords:** Anemia, HIV, Health-Related quality of life, AIDS

## Abstract

Anemia is a prevalent hematological complication in HIV patients, significantly impacting their health-related quality of life, morbidity, disease progression, and mortality. Despite its importance, there is a lack of information specifically focused on the impact of anemia on health-related quality of life (HRQoL) of HIV-infected individuals in Meru County, Kenya. This cross-sectional study aimed to determine the prevalence of anemia, identifying its predisposing factors, and assess its impact on HRQoL among HIV-infected individuals attending Meru Teaching and Referral Hospital (MeTRH).

A total of 354 adult participants (76.1% women, 24.9% men) were enrolled for the study at MeTRH between May and August 2022. Demographic and clinical data were collected using a standardized questionnaire. Hemoglobin levels were analyzed using an automated hemoglobin analyzer and anemia was defined according to WHO criteria (hemoglobin <11.5 g/dL for non-pregnant women, <12.5 g/dL for men). HRQoL was assessed using the WHOQOL-BREF tool across physical, psychological, social, and environment domains. To identify factors associated with anemia, logistic regression was used. Linear regression was applied to analyze the relationship between HRQoL mean scores and anemia status.

The prevalence of anemia in the study population was 12.4%, 11.4% in men and 12.8% in women. The level of education was the only sociodemographic factor found to be significantly associated with anemia status. Among the clinical factors, period of HIV positivity and CD4 count were found to be significantly associated with anemia status. The mean HRQOL score of the physical domain was statistically significantly different between the anemic and non-anemic groups. Furthermore, we found that anemia status was significantly associated with the physical domain of HRQoL after adjusting for period of HIV positivity, level of education and CD4 count (p = 0.02).

Anemia impacts the physical HRQoL in HIV patients, emphasizing the importance of comprehensive management strategies. Implementing a holistic approach to enhance physical well-being for these individuals is crucial in Meru County and similar resource-limited settings.

## INTRODUCTION

HIV remains a significant global health problem, with sub-Saharan Africa bearing the highest burden. In 2022, an estimated 39 million people worldwide were living with HIV (UNAIDS, 2023). By the end of 2022, Southern and Eastern Africa had an estimated 20.8 million HIV-positive individuals (UNAIDS, 2023). Although a decline in incidence of HIV infection has been observed, the number of people living with HIV (PLWHIV) is at an all-time high. This is due to increased access to antiretroviral therapy that has improved survival (Kharsany & Karim, 2016). Hematological complications have been found to be a significant problem among PLWHIV worldwide, negatively impacting their quality of life and disease progression (Melese et al., 2017; Cao et al., 2022a; Getu et al., 2023; Sah et al., 2020; Sahle et al., 2017). In Africa, the prevalence of anemia in PLWHIV ranges between 16.2% to 69% (Ikolango et al., 2023; Zenebe et al., 2019). The main causes of anemia in PLWHIV include nutrient deficiencies, anorexia, medication-related gastrointestinal disturbances, wasting, and malabsorption (Gardner & Kassebaum, 2020; Geletaw et al., 2017; Kumar et al., 2022). Anemia affects the overall functional status and well-being of PLWHIV, leading to symptoms such as fatigue, vertigo, dyspnea, and cardiovascular problems (Hoshino et al., 2020; Melese et al., 2017; Zuñiga et al., 2020).

The introduction of antiretroviral treatment (ART) significantly changed the trajectory of HIV, transforming it from a rapidly progressive illness to a chronic disease with reduced mortality and fewer opportunistic infections (Aynalem et al., 2020; Harding et al., 2020; Katemba et al., 2018; Melese et al., 2017). However, there has been insufficient attention to other health aspects such as emotional, psychological, financial, and physical well-being that may result in poor quality of life for many patients’ life (Algaralleh et al., 2020; Tesemma et al., 2019; Zenebe et al., 2019; Zou et al., 2022). Health-related quality of life encompasses health status, functional status, psychological well-being, and overall life satisfaction (Algaralleh et al., 2020; Osei-Yeboah et al., 2017; Tesemma et al., 2019; WHO, 2016).

There is limited research, specifically focused on Meru County, hindering the understanding of the local dynamics of HIV and anemia and their impact on the quality of life of PLWHIV. By addressing this gap, tailored interventions and strategies can be developed to improve outcomes for HIV positive patients in Meru County. Therefore, there is a pressing need for in-depth research to inform targeted interventions to address anemia among HIV positive individuals, particularly in underserved regions, to enhance their overall well-being and quality of life. This study investigated the association of anemia with health-related quality of life (HRQoL) in PLWHIV in Meru County. The four aspects of HRQoL including physical, social, environment and psychological were assessed.

## METHODOLOGY

### Study population

The cross-sectional study involved adult HIV positive patients attending Meru Teaching and Referral Hospital (MeTRH) in Meru, Kenya, between May and August 2022. A total of 354 HIV-Infected adults older than 18 years were recruited from the HIV Clinic at MeTRH. Ethical approval for the study was granted by Meru University Institutional Research Ethics Review Committee (MIRERC) and the MeTRH Ethics Review Committee. The eligibility criteria for participants for the study were; HIV positive adults aged 18 years and above consenting to participate in the study. Pregnant HIV positive women, children below 18 years and HIV positive adults who had been recently transfused (less than 6 months) were not eligible to take part in this study.

### Data collection

Questionnaires were administered to participants to collect socio-demographic data including age, sex, marital status, employment, residence, income and education level. Medical records from the MeTRH HIV clinic provided clinical information including HIV status, period under ART regimen, period of HIV positivity, CD4 level, TB diagnosis, WHO stage of HIV infection. World Health Organization Quality of Life BREF (WHOQOL-BREF) questionnaire, a shorter version of WHOQOL-100, was also administered to assess the health-related quality of life of the respondents under physical health, psychological health, social relationships and environment health domains (WHO, 2012). Physical health domain contains seven items that explore sleep, rest, pain, discomfort, energy and fatigue. Psychological health domain contains six items evaluating individuals’ perception of self-esteem, body image and appearance, positive and negative feelings, memory, learning, concentration and thinking. The social relations domain encompasses three items, social support, personal relationships and sexual activity. The environment domain entails eight items assessing safety and security, finances, health and social services access, home and physical environment and activities including education, training and leisure. Each item is rated on a 5-point Likert scale with a low score of one and a high score of five. To calculate the mean score for each domain, the mean score of the items for each domain is determined and then multiplied by 4 to make them comparable to WHOQOL-100. The domain mean scores are then transformed to a 0-100 scale by the formula (mean domain score-4) x (100/16).

Venous blood was drawn from the participants by a trained licensed lab technologist and placed in EDTA tubes. Hemoglobin levels were measured using an automated hematology analyzer. Quality control measures were implemented. Anemia was defined according to WHO criteria (hemoglobin <11.5 g/dL for non-pregnant women, <12.5 g/dL for men).

### Statistical analysis

Descriptive statistics was used to summarize the characteristics of the study population including socio-demographic and clinical information. Categorical data were represented by absolute and relative frequencies while continuous variables were represented by mean and standard deviation. A comparison of factors between anemia and non-anemic groups was performed using Pearson Chi square test, Z test and univarible logistic regression. Multivariable linear regression was performed to assess the relationship between anemia status and HRQoL mean score for each of the four domains and adjusted for potential confounders. The analysis was performed using R version 3.6.3 (R Core Team, 2021). The level of significance was set at p value <0.05 to determine statistical significance.

## RESULTS

### General Characteristics of the Study Population

A total of 354 HIV positive individuals, comprising of 266 (75.1%) females and 88 (24.9%) males were recruited for the study. The mean age of the participants was 44.7 years with a range between 20-77 years (Table 1). Majority of the participants (55.1%) had primary education being the highest level of education (Table 1). The mean hemoglobin level in the study population was 16.2 g/dL (6-29.9g/dL), with a mean of 16.4 g/dL (7.7-27.9 g/dL) in men and 16.1 g/dL (6-29.9g/dL) in women (Figure 1). The prevalence of anemia in study population was 12.4%, with 11.4% prevalence in men and 12.8% in women.

**Table 1:**
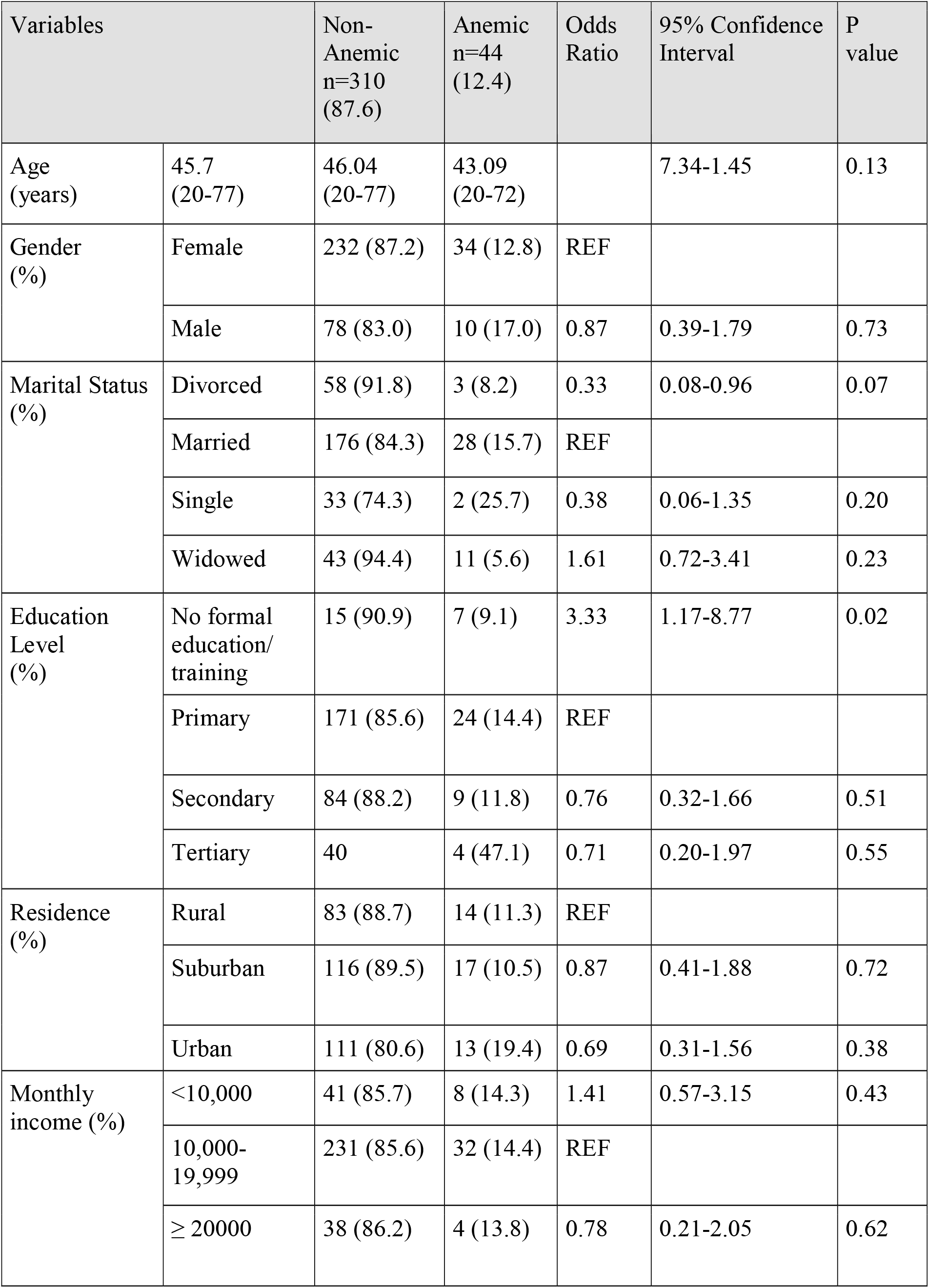
Socio-demographic characteristics according to anemia status.

| Variables |  | Non-Anemic<br>n=310<br>(87.6) | Anemic<br>n=44<br>(12.4) | Odds Ratio | 95% Confidence Interval | P value |
| --- | --- | --- | --- | --- | --- | --- |
| Age (years) | 45.7 (20-77) | 46.04 (20-77) | 43.09 (20-72) |  | 7.34-1.45 | 0.13 |
| Gender (%) | Female | 232 (87.2) | 34 (12.8) | REF |  |  |
|  | Male | 78 (83.0) | 10 (17.0) | 0.87 | 0.39-1.79 | 0.73 |
| Marital Status (%) | Divorced | 58 (91.8) | 3 (8.2) | 0.33 | 0.08-0.96 | 0.07 |
|  | Married | 176 (84.3) | 28 (15.7) | REF |  |  |
|  | Single | 33 (74.3) | 2 (25.7) | 0.38 | 0.06-1.35 | 0.20 |
|  | Widowed | 43 (94.4) | 11 (5.6) | 1.61 | 0.72-3.41 | 0.23 |
| Education Level (%) | No formal education/<br>training | 15 (90.9) | 7 (9.1) | 3.33 | 1.17-8.77 | 0.02 |
|  | Primary | 171 (85.6) | 24 (14.4) | REF |  |  |
|  | Secondary | 84 (88.2) | 9 (11.8) | 0.76 | 0.32-1.66 | 0.51 |
|  | Tertiary | 40 | 4 (47.1) | 0.71 | 0.20-1.97 | 0.55 |
| Residence (%) | Rural | 83 (88.7) | 14 (11.3) | REF |  |  |
|  | Suburban | 116 (89.5) | 17 (10.5) | 0.87 | 0.41-1.88 | 0.72 |
|  | Urban | 111 (80.6) | 13 (19.4) | 0.69 | 0.31-1.56 | 0.38 |
| Monthly income (%) | <10,000 | 41 (85.7) | 8 (14.3) | 1.41 | 0.57-3.15 | 0.43 |
|  | 10,000-19,999 | 231 (85.6) | 32 (14.4) | REF |  |  |
|  | ≥ 20000 | 38 (86.2) | 4 (13.8) | 0.78 | 0.21-2.05 | 0.62 |

**Figure 1:**
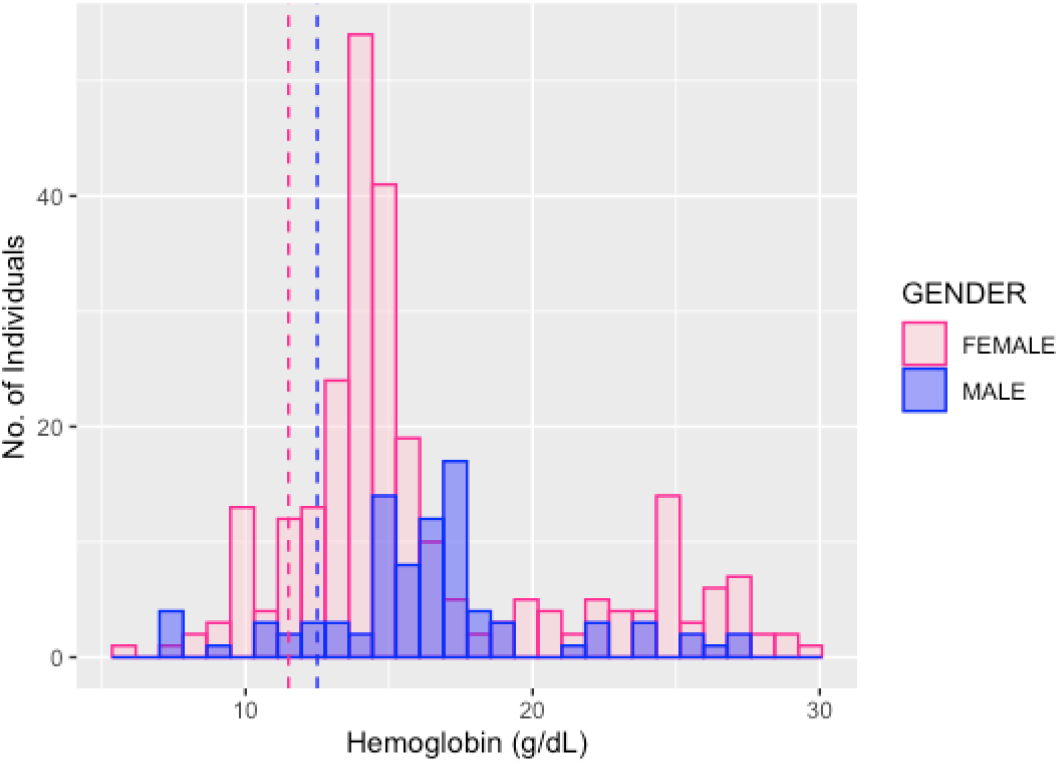
Distribution of hemoglobin level in the study population. Frequency distribution of hemoglobin levels in men indicated by blue bars and the distribution in female indicated by pink bars. Anemia was defined by hemoglobin level cutoffs indicated by vertical dashed lines at 11.5g/dL (pink) and 12.5g. dL (blue) in non-pregnant women and men respectively.

Among the socio-demographic factors assessed, we observed statistically significant association between the level of education with anemia status (univariate logistic regression, p value =0.02). The odds of anemia in individuals with no education and training was found to be 3.33 times higher compared to those with primary level of education (crude OR=3.33) (Table 1). Age, gender, marital status, residence and monthly income were not significantly associated with anemia status (p value > 0.05) (Table 1). Among the clinical factors assessed, CD4 count showed statistically significant association with anemia status (logistic regression, p value=0.002). Individuals with CD4 of greater than 400 c/mL had 0.35 decreased odds of being anemic as compared to individuals with CD4 levels less than 400 c/mL (Table 2). Other clinical characteristics including period of HIV positivity, WHO stage, TB diagnosis and period under ART treatment were not significantly associated with anemia status (p value > 0.05) (Table 2).

**Table 2:** Clinical characteristic according to anemia status.

| Variables |  | Anemia |  |  |  |  |
| --- | --- | --- | --- | --- | --- | --- |
|  |  | No<br>(n=310)<br>(87.6%) | Yes<br>(n=44)<br>(12.4 %) | Odd<br>Ratio | 95% CI | P value |
| Period of HIV positivity | ≤1 | 11 (68.8) | 5 (31.2) | 4.84 | 1.02-26.98 | 0.05 |
|  | >1 - 5 | 32 | 3 | REF |  |  |
|  | >5-10 | 112 | 16 | 1.52 | 0.47-6.84 | 0.52 |
|  | >10 | 155 | 20 | 1.38 | 0.44-6.09 | 0.78 |
| WHO Stage | I | 209 | 26 | REF |  |  |
|  | II | 99 | 16 | 1.30 | 0.66-2.51 | 0.44 |
| CD4 Count (c/ml) | ≤400 | 79 | 22 | REF |  |  |
|  | >400 | 215 | 21 | 0.35 | 0.18-0.67 | **0.002** |
| TB Diagnosis | Positive | 61 | 9 | 1.05 | 0.45-2.21 | 0.90 |
|  | Negative | 249 | 35 | REF |  |  |
| Period under ART treatment (Years) | 0 | 3 | 1 | 2.38 | 0.11-19.98 | 0.46 |
|  | >1-5 | 36 | 4 | 0.79 | 0.22-2.35 | 0.69 |
|  | >0-1 | 14 | 4 | 2.04 | 0.53-6.60 | 0.25 |
|  | ≥10 | 150 | 20 | 0.95 | 0.47-1.97 | 0.89 |
|  | >5-10 | 107 | 15 | REF |  |  |

In general, the non-anemic group had higher mean HRQoL scores compared to anemic group in all domains except the social relations domain (Table 3). Some participants reported very low HRQoL mean scores of less than 20 in the social relations domain (Figure 2). We observed that only the physical health domain mean HRQoL scores significantly differed between anemic and non-anemic groups (p-value= 0.004) (Table 3, Figure 2). No significant difference in mean scores between the groups was observed in psychological health, environment and social relation domains (p value >0.05) (Table 3, Figure 2).

**Table 3:** Comparison of HRQoL mean scores between anemic and non-anemic group.

| Group | Mean scores of HRQoL domains |  |  |  |
| --- | --- | --- | --- | --- |
|  | Physical Health | Psychological Health | Social Relations | Environment |
| Anemic mean(sd) | 53.08±7.69 | 55.02±9.10 | 50.37±18.93 | 47.72±13.01 |
| Non-Anemic mean(sd) | 56.75±7.14 | 55.25±9.26 | 48.65±18.26 | 49.78±11.79 |
| P value | **0.004** | 0.87 | 0.57 | 0.33 |

**Figure 2:**
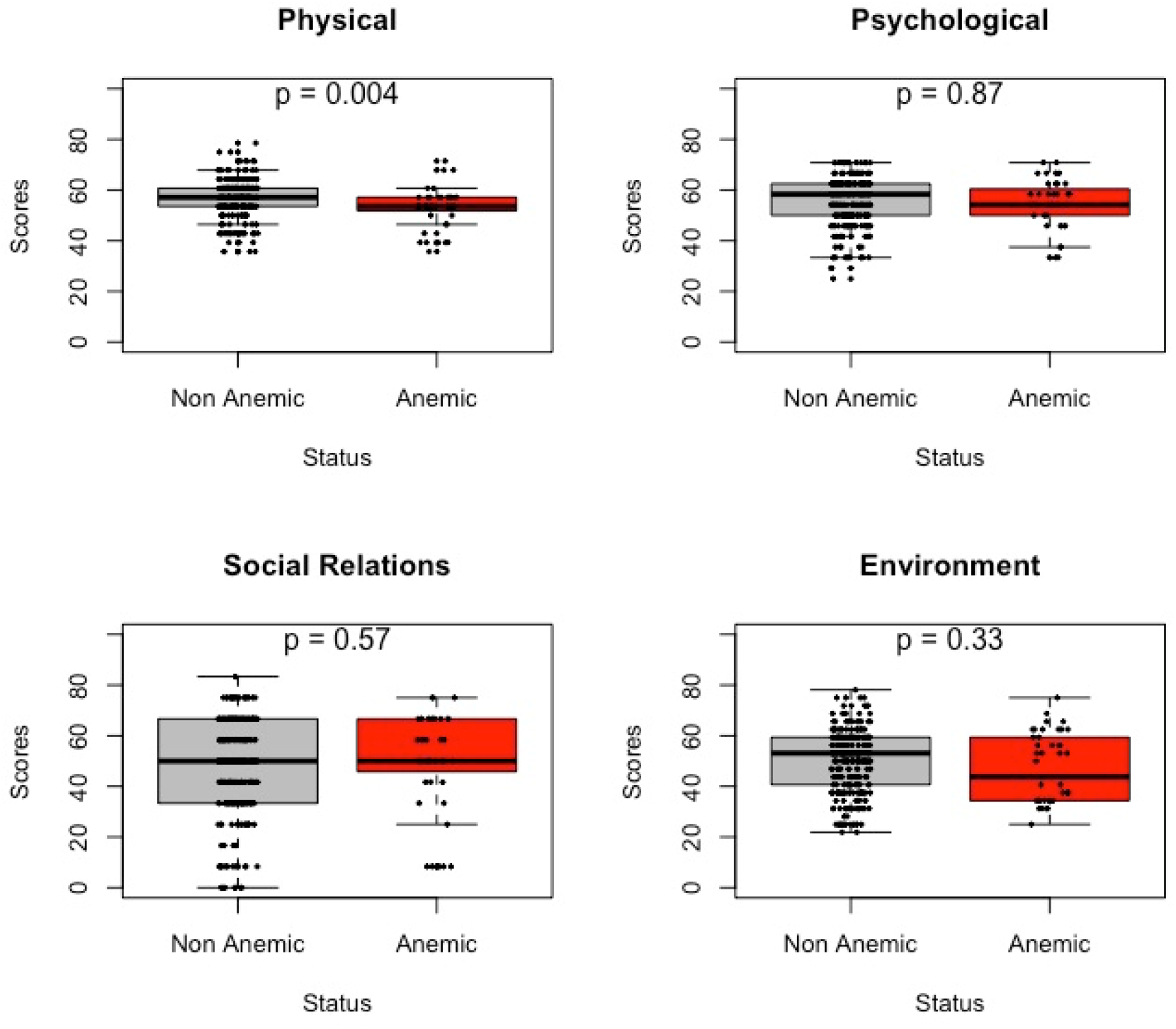
Distribution of mean HR-QoL scores of participants by anemia status. Box plot showing distribution of meant HR-QoL scores of anemic participants in black anemic participants in red under the physical health (top left), psychological health (top right), social relations (bottom left) and environment (bottom right) domains. P values indicated within each plot.

We further investigated factors, other than anemia, that may be associated with HRQoL. The period of HIV positivity was statistically significantly associated with physical and psychological health domains, education level was significantly associated with psychological and environment domains, and CD4 level was significantly associated with all domains except physical domain (univariable linear regression, p value <0.05) (Table 4). These significantly associated factors were further adjusted for in multivariable logistic regression analyzing the association between anemia status with each specific health domain.

**Table 4:** Socio-demographic and clinical factors other anemia showing significant association with HRQoL mean scores for the four domains by univariable linear regression analysis.

| Variable |  | Domain | Coef | 95% CI | P value |
| --- | --- | --- | --- | --- | --- |
| Period of HIV positivity in years | ≤1 | Physical | -0.60 | -0.12-0.09 | 0.09 |
|  | >1 - 5 |  | REF |  |  |
|  | >5-10 |  | -0.55 | -0.98-4.16 | **0.01** |
|  | >10 |  | -0.44 | -0.87- -0.22 | **0.04** |
| Level of education | No Education | Psychological | -0.005 | 0.53-1.87 | 0.99 |
|  | Primary |  | REF |  |  |
|  | Secondary |  | 0.58 | 1.25-2.55 | **0.001** |
|  | Tertiary |  | 1.07 | 1.83-4.66 | **<0.001** |
| Period of HIV positivity (years) | ≤1 | Psychological | -1.00 | -1.53- -0.46 | **<0.0001** |
|  | >1 - 5 |  | REF |  |  |
|  | >5-10 | Psychological | -1.00 | -1.53- -0.46 | **<0.0001** |
|  | >10 |  | -0.54 | -1.06- -0.02 | **0.04** |
| CD4 count (c/ml) | ≤400 |  | REF |  |  |
|  | >400 | Psychological | -1.20 | -1.52- -0.89 | **<0.001** |
|  | >400 | Social relations | -2.04 | -2.68- -1.39 | **<0.001** |
|  | >400 | Environment | -1.36 | -1.77- -0.95 | **<0.001** |
| Level of education | No Education | Environment | -0.55 | -1.38 - 0.28 | 0.20 |
|  | Primary |  | REF |  |  |
|  | Secondary |  | 0.13 | -0.34- 0.60 | **0.58** |
|  | Tertiary |  | 1.00 | 0.37 - 1.61 | **0.002** |

Anemia status was observed to be statistically significantly associated with physical health domain after adjusting for period of HIV positivity, level of education and CD4 count (multivariate logistic regression, p value = 0.02) (Table 5). The study revealed that individuals with anemia had a lower physical health HRQoL mean score of 2.80, on average, compared to non-anemia individual, adjusted for period of HIV positivity, level of education and CD4 count (Table 5). Psychological health, social relations and environmental domains were not significantly associated with HRQoL mean scores.

**Table 5:** Multivariate analysis of association between anemia status and HR-QoL mean score for the four domains adjusting for period of HIV positivity, level of education and CD4 count.

| **Domain** | **Coefficient** | **95% CI** | **P value** |
| --- | --- | --- | --- |
| Physical Health | -2.80 | -5.16 - 0.43 | **0.02** |
| Psychological Health | -0.90 | -3.56 - 1.76 | 0.51 |
| Social Relations | -2.10 | -7.87 - 3.67 | 0.48 |
| Environmental | -3.57 | -7.13 -0.02 | 0.05 |

## DISCUSSION

Anemia, a condition characterized by a decrease in the number of red blood cells or a reduction in hemoglobin levels, is a prevalent global health concern that impacts millions of individuals across various age groups (Cao et al., 2022b; Fekene et al., 2018; Hoshino et al., 2020; Katemba et al., 2018). While the physical consequences of anemia, such as fatigue, weakness, and impaired oxygen delivery, are well-documented, its broader implications on the overall health-related quality of life have gained increasing attention in recent years (Aksu & Ünal, 2023; Zuñiga et al., 2020).The association between anemia and health-related quality of life (HRQoL) has become a focal point of research, as it holds the potential to uncover significant insights into the multifaceted impacts of this condition on individuals’ well-being (Akbarpour et al., 2022; Durandt et al., 2019). Our study provides an in-depth analysis of the prevalence and determinants of anemia among HIV-positive individuals, as well as the impact of anemia on health-related quality of life (HRQoL). The findings are significant, especially in the case of resource-limited settings such as Kenya, where both HIV and anemia pose considerable public health challenges.

The overall prevalence of anemia in our study population was 12.4%, with a slightly higher prevalence in women (12.8%) compared to men (11.4%), which contrasts with the higher prevalence rates reported in several other studies. For instance, Melese et al. (2017) reported a prevalence of 23% in Debre-Tabor Hospital, Ethiopia, and Wendie and Mengistu (2022) found a prevalence of 41.3% among HIV-positive patients. The study by Sah et al. (2020) found an even higher prevalence of 66.7% in Nepal, indicating significant geographic and demographic variability (Aynalem et al., 2020; Melese et al., 2017; Wendie & Mengistu, 2022). This variability can be attributed to several factors, including differences in study populations, methodologies, healthcare settings, and the criteria used to define anemia.

Level of education emerged as a significant socio-demographic factor associated with anemia status in our study. The role of education in anemia among HIV-positive patients is notable. Studies such as Gebremedhin & Haye (2019) and Zenebe et al. (2019) have reported that lower education levels are associated with a higher risk of anemia. Individuals with lower educational attainment may have limited access to health information, which can affect their understanding of anemia management and nutritional needs. This is supported by the finding from Zenebe et al. (2019) that inability to read and write was a significant predictor of anemia, suggesting that educational interventions could play a crucial role in anemia prevention and management (Gebremedhin & Haye, 2019; Zenebe et al., 2019).

Among clinical factors, CD4 count was significantly associated with anemia. Individuals with CD4 counts greater than 400 cells/mL had a lower likelihood (OR = 0.35) of being anemic compared to non-anemic. Studies consistently show that as CD4 count decreases, the prevalence of anemia increases. Sah et al. (2020) reported anemia prevalence rates of 5.71%, 12.85%, and 48.09% among patients with CD4 counts >500, 200–499, and <200 cells/mm^3^, respectively. This relationship highlights the importance of maintaining adequate immune function to mitigate the risk of anemia (Sah et al., 2020). However, other clinical variables such as the duration of HIV positivity, WHO stage, TB diagnosis, and duration of ART did not show a significant association with anemia status in our study.

Our investigation into the relationship between anemia and HRQoL revealed that anemia significantly impacts the physical health domain. Anemic individuals reported lower physical health HRQoL mean scores compared to their non-anemic counterparts, after adjusting for period of HIV positivity, education level, and CD4 count. This finding is critical as it highlights the tangible impact of anemia on daily functioning and overall well-being of HIV-positive individuals.

Interestingly, while anemia was significantly associated with physical health, it did not show a significant relationship with psychological, social, or environmental domains of HRQoL. This suggests that the physical manifestations of anemia, such as fatigue and reduced physical capacity, are more immediately perceptible to patients compared to its effects on other aspects of life. This fundings correspond to the findings of studies carried out in different settings. A study carried out in US and Brazil is consistent with our study and indicates the role anemia has on the health-related quality of life (Hoshino et al., 2020). Another study carried out in Ethiopia also concurred with the findings of our study which stated that risk factors such as low CD4 count, opportunistic infections and anemia contribute negatively to the health-related quality of life among HIV positive patients (Osei-Yeboah et al., 2017).

The associations between anemia, education level, and CD4 count emphasize the need for integrated care approaches that address both clinical and socio-economic determinants of health. Routine screening for anemia in HIV-positive individuals, particularly those with lower CD4 counts or lower educational attainment, should be prioritized. Furthermore, interventions aimed at improving educational opportunities and health literacy could play a role in reducing the burden of anemia. The significant impact of anemia on physical HRQoL emphasizes the importance of early detection and management of anemia to improve overall health outcomes and quality of life in HIV-positive populations. This could involve nutritional interventions, iron supplementation, or adjustments in ART regimens to minimize hematologic toxicity.

Despite the robust findings, our study has some limitations. The cross-sectional design limits the ability to infer causality between anemia and HRQoL. Longitudinal studies are needed to further explore these associations over time. Additionally, our study was conducted in a specific geographical and socio-economic context, which may limit the generalizability of the findings to other settings. Future research should aim to identify specific causes of anemia in HIV-positive individuals, such as nutritional deficiencies, chronic infections, or ART-related side effects. Investigating these factors could provide more targeted strategies for anemia prevention and management.

## Data Availability

All data produced in the present study are available upon reasonable request to the authors

